# On the Analysis of Mortality Risk Factors for Hospitalized COVID-19 Patients: a Data-driven Study Using the Major Brazilian Database

**DOI:** 10.1101/2020.09.24.20200766

**Authors:** Fernanda Sumika Hojo de Souza, Natália Satchiko Hojo-Souza, Ben Dêivide de Oliveira Batista, Cristiano Maciel da Silva, Daniel Ludovico Guidoni

## Abstract

**Background:** Brazil became the epicenter of the COVID-19 epidemic in a brief period of a few months after the first officially registered case. The knowledge of the epidemiological/clinical profile and the risk factors of Brazilian COVID-19 patients can assist in the decision making of physicians in the implementation of early and most appropriate measures for poor prognosis patients. However, these reports are missing. Here we present a comprehensive study that addresses this demand.

**Methods:** This data-driven study was based on the Brazilian Ministry of Health Database (SIVEP-Gripe, 2020) regarding notified cases of hospitalized COVID-19 patients during the period from February 26 to August 10, 2020. Demographic data, clinical symptoms, comorbidities and other additional information of patients were analyzed.

**Results:** The hospitalization rate was higher for male gender (56.56%) and for older age patients of both sexes. Overall, the mortality rate was quite high (41.28%) among hospitalized patients, especially those over 60 years of age. Most prevalent symptoms were cough, dyspnoea, fever, low oxygen saturation and respiratory distress. Heart disease, diabetes, obesity, kidney disease, neurological disease, and pneumopathy were the most prevalent comorbidities. A high prevalence of hospitalized COVID-19 patients with heart disease (65.7%) and diabetes (53.55%) and with a high mortality rate of around 50% was observed. The ICU admission rate was 39.37% and of these 62.4% died. 24.4% of patients required invasive mechanical ventilation (IMV), with high mortality among them (82.98%). The main mortality risk predictors were older age and IMV requirement. In addition, socioeconomic conditions have been shown to significantly influence the disease outcome, regardless of age and comorbidities.

**Conclusion:** Our study provides a comprehensive overview of the hospitalized Brazilian COVID-19 patients profile and the mortality risk factors. The analysis also evidenced that the disease outcome is influenced by multiple factors, as unequally affects different segments of population.

## 1. INTRODUCTION

A new highly infectious-contagious coronavirus for humans emerged in December 2019 in Wuhan (Hubei province, China) [1], [2]. The initial outbreak in the Chinese region spread rapidly, but only on March 11, 2020 it was recognized as a pandemic by WHO [3]. The disease, called COVID-19, can cause severe pneumonia due to SARS-CoV-2 (severe acute respiratory syndrome coronavirus 2) infection. However, compared to other respiratory coronavirus syndromes (SARS-CoV and MERS-CoV), the mortality rate is low despite the high potential for spread [4].

The worldwide traffic of people facilitated the rapid spread of COVID-19, reaching countries on all continents in a short time. According to WHO data from September 15, 2020, more than 29 million cases of COVID-19 have been confirmed, including over 920,000 deaths worldwide. Brazil has officially recorded the first case of COVID-19 on February 26, 2020, and has become the epicenter of the pandemic with over 4.3 million confirmed COVID-19 cases and more than 131,000 deaths on September 15, 2020 [5].

COVID-19 has a broad spectrum of the disease, ranging from asymptomatic to extremely severe cases. According to WHO, the majority (about 80%) of COVID-19 patients is asymptomatic or mild, 15% are severe, requiring oxygen, and 5% are critical cases demanding mechanical ventilation [5]. However, recent studies suggest that asymptomatic individuals are 40 to 45% of the infected population [6] or even only 20% [7].

The symptomatic infection is characterized by fever, generalized weakness, dry cough, headache, dyspnoea, and myalgia, as well as leukopenia, lymphocytopenia, neutrophilia, high levels of C-reactive protein, D-dimer, lactate dehydrogenase and inflammatory cytokines [8], [2], [9] and loss of smell and taste in the initial stage of infection [10]. Although biomarkers such as lactate dehydrogenase, D-dimer and inflammatory cytokines, as well as chest computed tomography are good disease indicators, these tests are not routinely performed in most health centers. Therefore, in clinical practice, this parameter set does not help in the diagnosis and prognosis of the affected population, mainly in developing countries.

Symptomatic cases of COVID-19 may progress to recovery or a very severe condition, characterized by acute respiratory distress syndrome (ARDS), cytokine storm, blood coagulation dysfunction, acute cardiac and kidney injury, and multi-organ dysfunction, resulting in patient death [1], [11], [12], [13]. Age and comorbidities may influence the disease outcome. In fact, several studies have shown that the elderly and the presence of comorbidities such as heart disease, diabetes, chronic lung disease and obesity contribute to a more severe infection outcome [9], [14].

There is still no vaccine or therapeutic drugs for the specific treatment of COVID-19. Therefore, quarantine and social distancing are being the recommended measures in an attempt to reduce the infection rate in order to avoid overloading healthcare systems [15].

In Brazil, it is believed that the health system will collapse for patients with COVID-19 due to the lack of beds, ventilators in ICUs and Personal Protective Equipment (PPE) by health professionals given the growing number of cases beyond seasonal influenza cases. Bed occupancy in several Brazilian states is over 80%, at a time when the country reached the peak of the pandemic and continues on a high plateau. This is also true in many other countries. Thus, the screening of hospitalized patients with COVID-19 at risk of poor prognosis, requiring admission to the ICU and mechanical ventilation, could improve the flow of care in hospitals, avoid overload and contribute to reducing the mortality rate.

Data from unidentified COVID-19 positive patients who have been hospitalized are available on the integrated health surveillance platform of the Brazil Ministry of Health. However, the database, named Brazilian Severe Acute Respiratory Syndrome Database of the SIVEP Gripe^1^ (Sistema de Informação de Vigilância Epidemiológica da Gripe) including data from COVID-19 in addition to Influenza, displays incomplete reports for demographic data, symptoms, comorbidities, ICU admission, etc.

A comprehensive study on the characteristics and risk factors of Brazilian COVID-19 patients is missing. Thus, herein, we propose:

- Building a dataset containing demographic data, clinical symptoms and comorbidities, ICU admission and use of mechanical ventilation of Brazilian hospitalized COVID-19 patients: those who were discharged and those who deceased, in order to investigate possible risk factors.
- Analyzing the mortality risk factors for different subgroups of hospitalized COVID-19 patients using the created dataset.

## 2. DATA COLLECTION AND METHODS

This retrospective study is based on a publicly available database and did not directly involve patients; it did not require approval by an ethics committee.

### 2.1. Dataset Creation

In order to build the dataset to outline the profiles of hospitalized COVID-19 patients, we downloaded data from the Brazilian Severe Acute Respiratory Syndrome Database (SIVEP-Gripe, 2020) covering the time from February 26 to August 10, 2020. As shown in Figure 1, 225,987 patients were positive SARS-COV-2 confirmed by RT-PCR method, with 208,969 patients being admitted to hospital according to information contained in SIVEP Database. We used in our study data from 162,045 patients who had closed outcome (cure or death) in order to provide a profile overview of the patients and after, a 44,128 patients cohort with full symptom/comorbidity information aiming to analyze risk factors for mortality.

**Figure 1:**
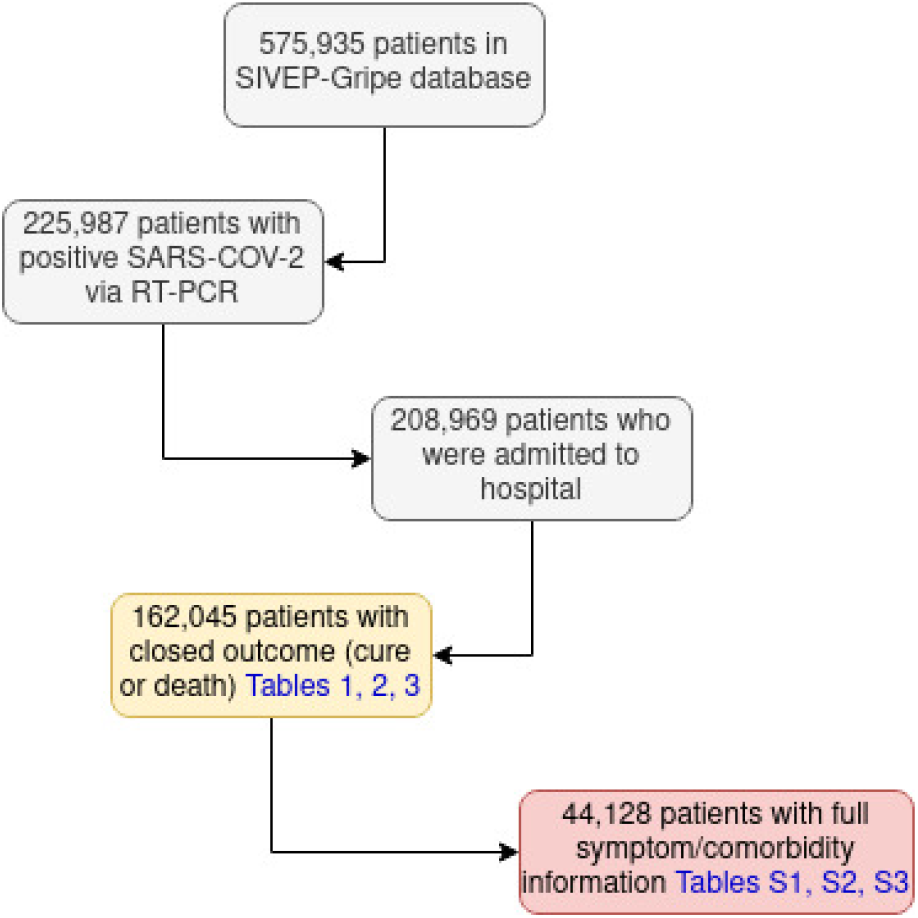
Flowchart of SIVEP-Gripe data used in the study

### 2.2. Statistical analysis

Categorical variables are given in absolute numbers and percentages. Continuous variables are reported through medians with IQRs (interquartile range). Survival curves were plotted using the Kaplan-Meier method and compared using a generalization of the log-rank test, which can deal with *n >* 2 populations. Cox proportional hazards model was used to estimate hazard ratios (HR) in simple and multiple regression models analysis. Variables in the adjusted models were selected following the strategy proposed by Collett [16], consisting of a step-wise, iterative, optimizing algorithm^2^ started with all variables with *p <* 0.10 on the simple regression analysis.

All analyses were performed using Python (version 3.6.9) and the statistical package lifelines (version 0.24.16) [17].

## 3. RESULTS

### 3.1. Profile of hospitalized COVID-19 patients

Our study dataset included information of hospitalized COVID-19 patients, confirmed by RT-PCR. Demographic data of the study population were disaggregated into the cure and death subgroups (Table 1). The overall mortality rate was 41.28%, with a predominance of cases in the Southeast region (58.96%) in the covered period. The number of cases was higher among the elderly over 60 years (53.20%), male gender (56.56%) and, white (50.25%) and brown color patients (41.03%).

**TABLE 1:** Demographic data of the study population

|  | all n(%) | cure n(%) | death n(%) | lr<sup>d</sup>(%) |
| --- | --- | --- | --- | --- |
| Region | 162045 (100.00) | 95149 (58.72) | 66896 (41.28) |  |
| South | 16088 (9.93) | 10878 (11.43) | 5210 (7.79) | 32.38 |
| Southeast | 95541 (58.96) | 60169 (63.24) | 35372 (52.88) | 37.02 |
| Midwest | 9928 (6.13) | 6227 (6.54) | 3701 (5.53) | 37.28 |
| Northeast | 30114 (18.58) | 13269 (13.95) | 16845 (25.18) | 55.94 |
| North | 10374 (6.40) | 4606 (4.84) | 5768 (8.62) | 55.60 |
| Age Range<sup>b</sup> |  |  |  |  |
| 0-4 | 1458 (0.90) | 1265 (1.33) | 193 (0.29) | 13.24 |
| 5-9 | 348 (0.21) | 311 (0.33) | 37 (0.06) | 10.63 |
| 10-19 | 999 (0.62) | 837 (0.88) | 162 (0.24) | 16.22 |
| 20-29 | 5312 (3.28) | 4598 (4.83) | 714 (1.07) | 13.44 |
| 30-39 | 15381 (9.49) | 13127 (13.80) | 2254 (3.37) | 14.65 |
| 40-49 | 22952 (14.16) | 18202 (19.13) | 4750 (7.10) | 20.70 |
| 50-59 | 29383 (18.13) | 20312 (21.35) | 9071 (13.56) | 30.87 |
| 60-69 | 32935 (20.32) | 17634 (18.53) | 15301 (22.87) | 46.46 |
| 70-79 | 28677 (17.70) | 11648 (12.24) | 17029 (25.46) | 59.38 |
| 80-89 | 19159 (11.82) | 5939 (6.24) | 13220 (19.76) | 69.00 |
| 90+ | 5441 (3.36) | 1276 (1.34) | 4165 (6.23) | 76.55 |
| Gender |  |  |  |  |
| male | 91656 (56.56) | 52874 (55.57) | 38782 (57.97) | 42.31 |
| female | 70389 (43.44) | 42275 (44.43) | 28114 (42.03) | 39.94 |
| Race | 110503 (100.00) | 63173 (57.17) | 47330 (42.83) |  |
| white | 55533 (50.25) | 34145 (54.05) | 21388 (45.19) | 38.51 |
| black | 7683 (6.95) | 4184 (6.62) | 3499 (7.39) | 45.54 |
| asian | 1659 (1.50) | 924 (1.46) | 735 (1.55) | 44.30 |
| brown | 45334 (41.03) | 23772 (37.63) | 21562 (45.56) | 47.56 |
| indigenous | 294 (0.27) | 148 (0.23) | 146 (0.31) | 49.66 |
| Education<sup>c</sup> | 54547 (100.00) | 33117 (60.71) | 21430 (39.29) |  |
| illiterate | 3321 (6.09) | 1156 (3.49) | 2165 (10.10) | 65.19 |
| ES-1 | 13767 (25.24) | 6560 (19.81) | 7207 (33.63) | 52.35 |
| ES-2 | 10114 (18.54) | 5705 (17.23) | 4409 (20.57) | 43.59 |
| HS | 17527 (32.13) | 12042 (36.36) | 5485 (25.59) | 31.29 |
| HE | 8722 (15.99) | 6710 (20.26) | 2012 (9.39) | 23.07 |
| NA | 1096 (2.01) | 944 (2.85) | 152 (0.71) | 13.87 |

a. lethality rate
b. in years
c. ES-1= Elementary School 1; ES-2= Elementary School 2; HS= High School; HE= Higher Education, NA= Not Applicable (age<7)

The distribution by sex and age group is shown in Figure 2. The mortality rate in hospitalized COVID-19 patients aged *<*20 years was very low, in accordance with observations made in other populations. The mortality rate was higher in the elderly of both sexes.

**Figure 2:**
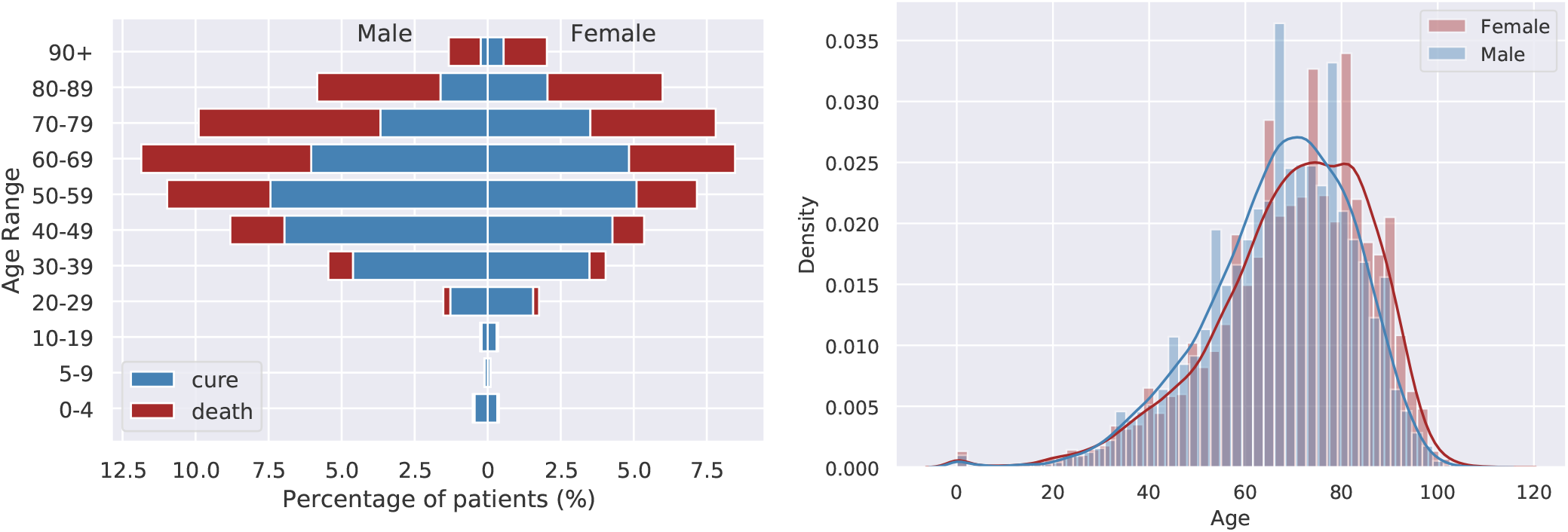
Outcome (a) and mortality density (b) distribution by age disaggregated data and gender of the study population (n=162,045)

Besides a higher hospitalization rate in male gender (56.56%), the mortality rate was significantly higher in males (57.97%) compared to females (42.03%) (*p <* 0.001) (Table 1). Moreover, age-disaggregated data (Figure 2) shows that the influence of gender on the risk of death is dependent on the age range.

Lethality rate analysis showed a high rate in the North/Northeast region, in the highest age groups, in non-white populations and in the lowest educational levels (see Table 1 and Figure 3). The higher mortality in the North/Northeast region may be due to socioeconomic conditions and availability of beds in the ICU. On the other hand, it may also be due to the lack of appropriate knowledge about the characteristics of the new disease whose spread was highest in this region at the beginning of the pandemic.

**Figure 3:**
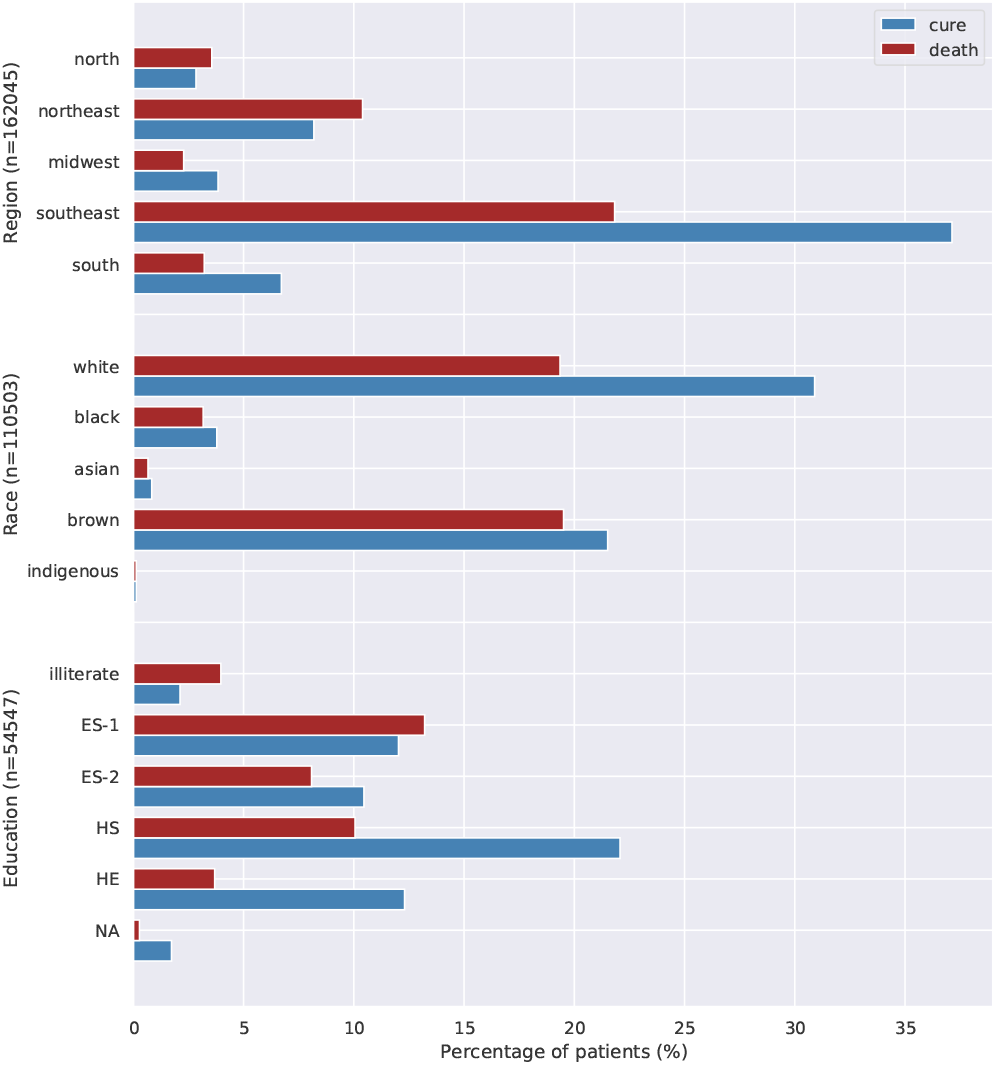
Outcome distribution according to demographic characteristics. Different number of patients apply (Brazilian regions: n=162,045, race: n=110,503, education: n=54,547).

The most frequent symptoms and comorbidities in hospitalized COVID-19 patients are shown in Figure 4 and Table 2. Importantly, cough, dyspnoea, fever, low oxygen saturation and respiratory distress (Figure 4a), and heart disease, diabetes, obesity, kidney disease, neuropathy, and pneumopathy (Figure 4b) were more prevalent.

**TABLE 2:** Clinical data of the study population

|  | all n(%) | cure n(%) | death n(%) |
| --- | --- | --- | --- |
| Symptom |  |  |  |
| Fever (n=145071) | 111435 (76.81) | 68783 (61.72) | 42652 (38.28) |
| Cough (n=146497) | 120292 (82.11) | 74154 (61.64) | 46138 (38.36) |
| Sore Throat (n=119158) | 29420 (24.69) | 19261 (65.47) | 10159 (34.53) |
| Dyspnoea (n=145342) | 116132 (79.90) | 64024 (55.13) | 52108 (44.87) |
| Respiratory Distress (n=134430) | 92991 (69.17) | 49453 (53.18) | 43538 (46.82) |
| SP O2 < 95%<sup>a</sup> (n=135790) | 95080 (70.02) | 48772 (51.30) | 46308 (48.70) |
| Diarrhea (n=117100) | 21536 (18.39) | 14367 (66.71) | 7169 (33.29) |
| Vomit (n=114281) | 12298 (10.76) | 7953 (64.67) | 4345 (35.33) |
| Other (n=116778) | 55974 (47.93) | 37771 (67.48) | 18203 (32.52) |
| Comorbidity |  |  |  |
| Cardiac disease (n=86244) | 56664 (65.70) | 28302 (49.95) | 28362 (50.05) |
| Hematological disease (n=63015) | 1544 (2.45) | 725 (46.96) | 819 (53.04) |
| Down's syndrome (n=62832) | 433 (0.69) | 211 (48.73) | 222 (51.27) |
| Liver disease (n=62711) | 1613 (2.57) | 635 (39.37) | 978 (60.63) |
| Asthma (n=63873) | 4566 (7.15) | 3047 (66.73) | 1519 (33.27) |
| Diabetes (n=80155) | 42919 (53.55) | 20824 (48.52) | 22095 (51.48) |
| Neuropathy (n=65159) | 7075 (10.86) | 2598 (36.72) | 4477 (63.28) |
| Pneumopathy (n=64721) | 6627 (10.24) | 2603 (39.28) | 4024 (60.72) |
| Immunodepression (n=63844) | 5195 (8.14) | 2370 (45.62) | 2825 (54.38) |
| Kidney disease (n=64884) | 7601 (11.71) | 2693 (35.43) | 4908 (64.57) |
| Obesity (n=63022) | 7413 (11.76) | 4196 (56.60) | 3217 (43.40) |
| Other (n=77316) | 45849 (59.30) | 23199 (50.60) | 22650 (49.40) |

a. blood oxygen saturation

**Figure 4:**
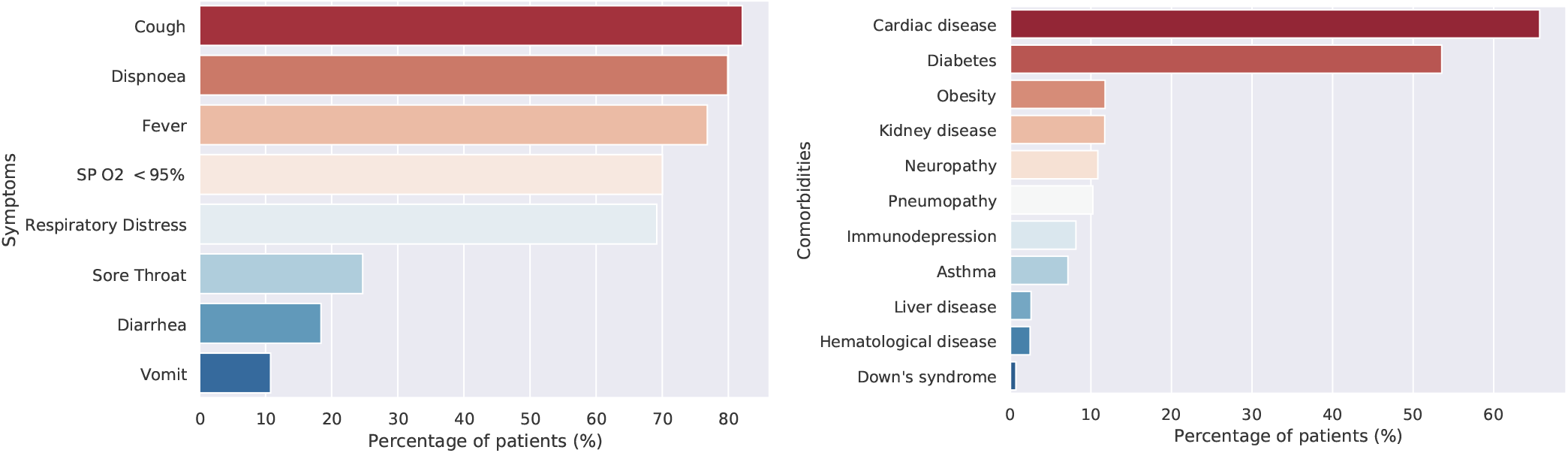
Clinical characteristics (a) and comorbidities (b) most frequent in the study population

Additional information from hospitalized COVID-19 patients is shown in Table 3. The ICU admission rate was 39.37% (of n=145,053), with 62.40% death. In addition, 24.41% (of n=138,962) required invasive mechanical ventilation, and of these 82.98% died. In contrast, only 31.83% died among those who required non-invasive ventilation. At least 72% (of n=138,962) required some kind of ventilation, indicating that hospitalized patients can lead to the collapse of healthcare systems. The mortality rate in the Influenza vaccinated group (33.64% of n=62.695) and in the Influenza antiviral group (37.92% of n=124,844) were 37.82% and 39.80%, respectively.

**TABLE 3:**
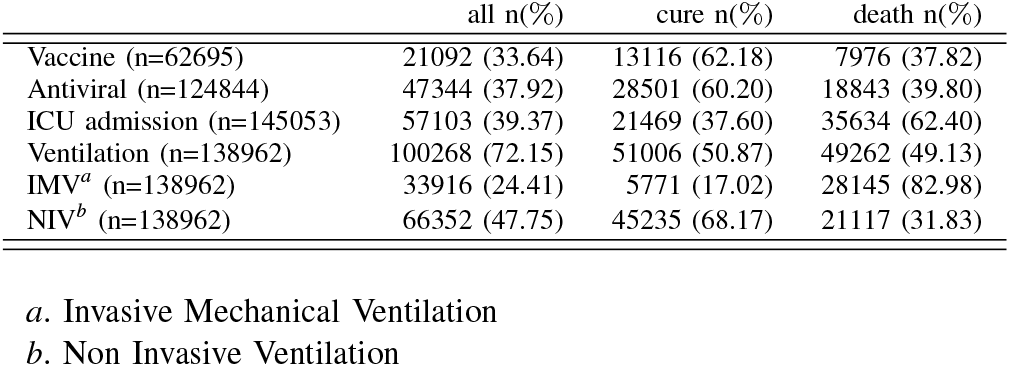
Additional information of the study population

|  | all n(%) | cure n(%) | death n(%) |
| --- | --- | --- | --- |
| Vaccine (n=62695) | 21092 (33.64) | 13116 (62.18) | 7976 (37.82) |
| Antiviral (n=124844) | 47344 (37.92) | 28501 (60.20) | 18843 (39.80) |
| ICU admission (n=145053) | 57103 (39.37) | 21469 (37.60) | 35634 (62.40) |
| Ventilation (n=138962) | 100268 (72.15) | 51006 (50.87) | 49262 (49.13) |
| IMV<sup>a</sup> (n=138962) | 33916 (24.41) | 5771 (17.02) | 28145 (82.98) |
| NIV<sup>b</sup> (n=138962) | 66352 (47.75) | 45235 (68.17) | 21117 (31.83) |

*a.* Invasive Mechanical Ventilation
*b.* Non Invasive Ventilation

### 3.2. Risk factors for mortality

Cox regression model was adapted for simple and multiple regression analyses, considering the time between hospitalization and outcome (median=8 days, IQR 4-14). Such analyzes were based only on patients with complete data (n=44,128) for symptoms, comorbidities, information regarding ICU admission, ventilation and temporal values, besides age and gender (Tables S1, S2 and S3). Vaccine and antiviral information were considered negative in the absence of data. The general characteristics of this subgroup are representative of the study population presented in Tables 1, 2 and 3.

Although the lethality rate is ≈3.5% for notified COVID-19 cases in Brazil, among hospitalized patients the condition is very critical, with a ≈41% lethality according to Table 1. Mortality HRs for hospitalized COVID-19 patients are shown in Figure 5.

**Figure 5:**
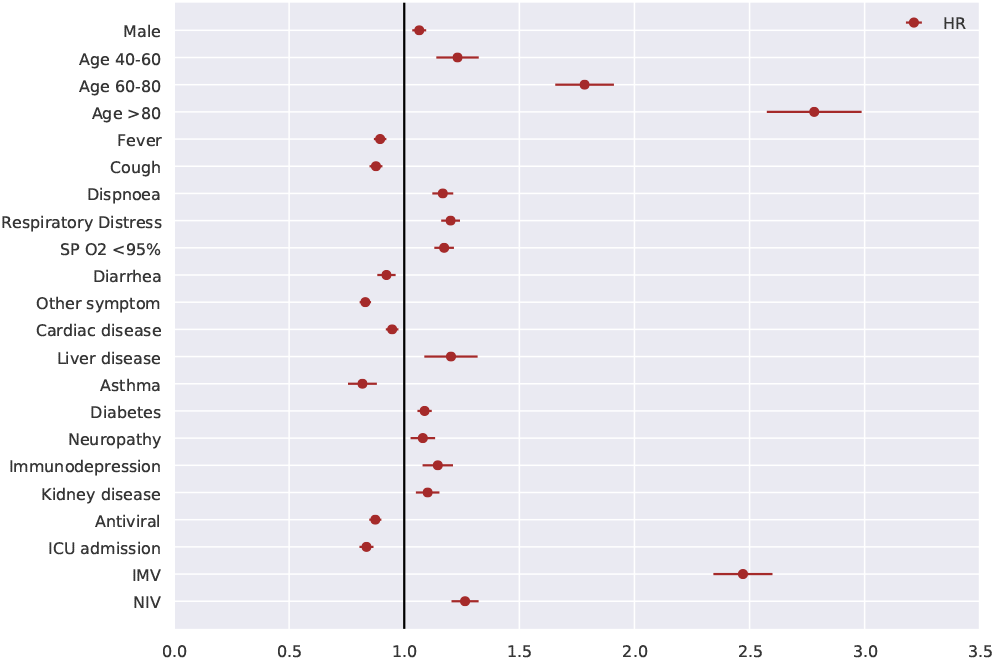
Mortality prognosis based on the general characteristics of patients in a multiple Cox regression model (CI 95%) n=44,128. (See Table 4)

The simple Cox regression analysis presented 26 variables^3^(out of 28) as potential increase/decrease risk factors for mortality (Table S4). However, after applying the variable selection strategy [16] for a multiple regression model, 20 variables remained statistically significant in the multiple regression analysis (Table 4), with a model concordance index of 0.69.

**TABLE 4:** Risk factors in fatal outcome using a multiple Cox regression model (95% CI)

| Variable<sup>a</sup> | HR | CI 95% | p value |
| --- | --- | --- | --- |
| Male | 1.07 | (1.04-1.10) | <0.001 |
| Age 40-60 | 1.23 | (1.14-1.33) | <0.001 |
| Age 60-80 | 1.78 | (1.66-1.92) | <0.001 |
| Age >80 | 2.78 | (2.58-2.99) | <0.001 |
| Fever | 0.90 | (0.87-0.92) | <0.001 |
| Cough | 0.88 | (0.85-0.91) | <0.001 |
| Dispnoea | 1.17 | (1.12-1.21) | <0.001 |
| Respiratory Distress | 1.20 | (1.16-1.24) | <0.001 |
| SP O2 <95% | 1.17 | (1.13-1.22) | <0.001 |
| Diarrhea | 0.92 | (0.88-0.96) | <0.001 |
| Other symptom | 0.83 | (0.81-0.86) | <0.001 |
| Cardiac disease | 0.95 | (0.92-0.98) | <0.001 |
| Liver disease | 1.20 | (1.09-1.32) | <0.001 |
| Asthma | 0.82 | (0.76-0.88) | <0.001 |
| Diabetes | 1.09 | (1.06-1.12) | <0.001 |
| Neuropathy | 1.08 | (1.03-1.14) | <0.005 |
| Immunodepression | 1.15 | (1.08-1.21) | <0.001 |
| Kidney disease | 1.10 | (1.05-1.15) | <0.001 |
| Antiviral | 0.87 | (0.85-0.90) | <0.001 |
| ICU admission | 0.84 | (0.81-0.87) | <0.001 |
| IMV | 2.47 | (2.35-2.60) | <0.001 |
| NIV | 1.26 | (1.21-1.32) | <0.001 |

<sup>a</sup>. see Table S2; n=44,128 patients with full symptom/comorbidity information

According to the multiple Cox regression analysis, older age: 40-60 years old (HR=1.23; 95% CI, 1.14-1.33), 60-80 years old (HR=1.78; 95% CI, 1.66-1.92), *>*80 years old (HR=2.78; 95% CI, 2.58-2.99) and invasive mechanical ventilation (HR=2.47; 95% CI, 2.35-2.60) were the main risk factors for mortality. Besides that, noninvasive ventilation (HR=1.26; 95% CI, 1.21-1.32), respiratory distress (HR=1.20; 95% CI, 1.16-1.24), liver disease (HR=1.20; 95% CI, 1.09-1.32), dyspnoea (HR=1.17; 95% CI, 1.12-1.21), low oxygen saturation (HR=1.17; 95% CI, 1.13-1.22),immunosuppression (HR=1.15; 95% CI, 1.08-1.21), kidney disease (HR=1.10; 95% CI, 1.05-1.15), diabetes (HR=1.09; 95% CI, 1.06-1.12), neuropathy (HR=1.08; 95% CI, 1.03-1.14) and male gender (HR=1.07; 95% CI, 1.04-1.10) were also significantly associated with poor prognosis (Figure 5, Table 4).

Importantly, asthma comorbidity was not a mortality risk factor. In addition, the use of Influenza antivirals (oseltamivir or zanamivir) showed a small reduction in the mortality risk (HR=0.87; 95% CI, 0.85-0.90), but Influenza vaccination showed no statistical significance in the multiple Cox regression analysis (Figure 5, Table 4).

It is interesting to note that ICU admission was not a risk factor for mortality, emphasizing that the most important feature is the invasive or non-invasive ventilation requirement. Kaplan-Meier survival curves for the most significant prognostic factors (older age and ventilation requirement) are shown in Figure 6. Ventilation requirement was a strong predictor of mortality risk factor, as shown in Figure 6a, as well as older age (Figure 6b).

**Figure 6:**
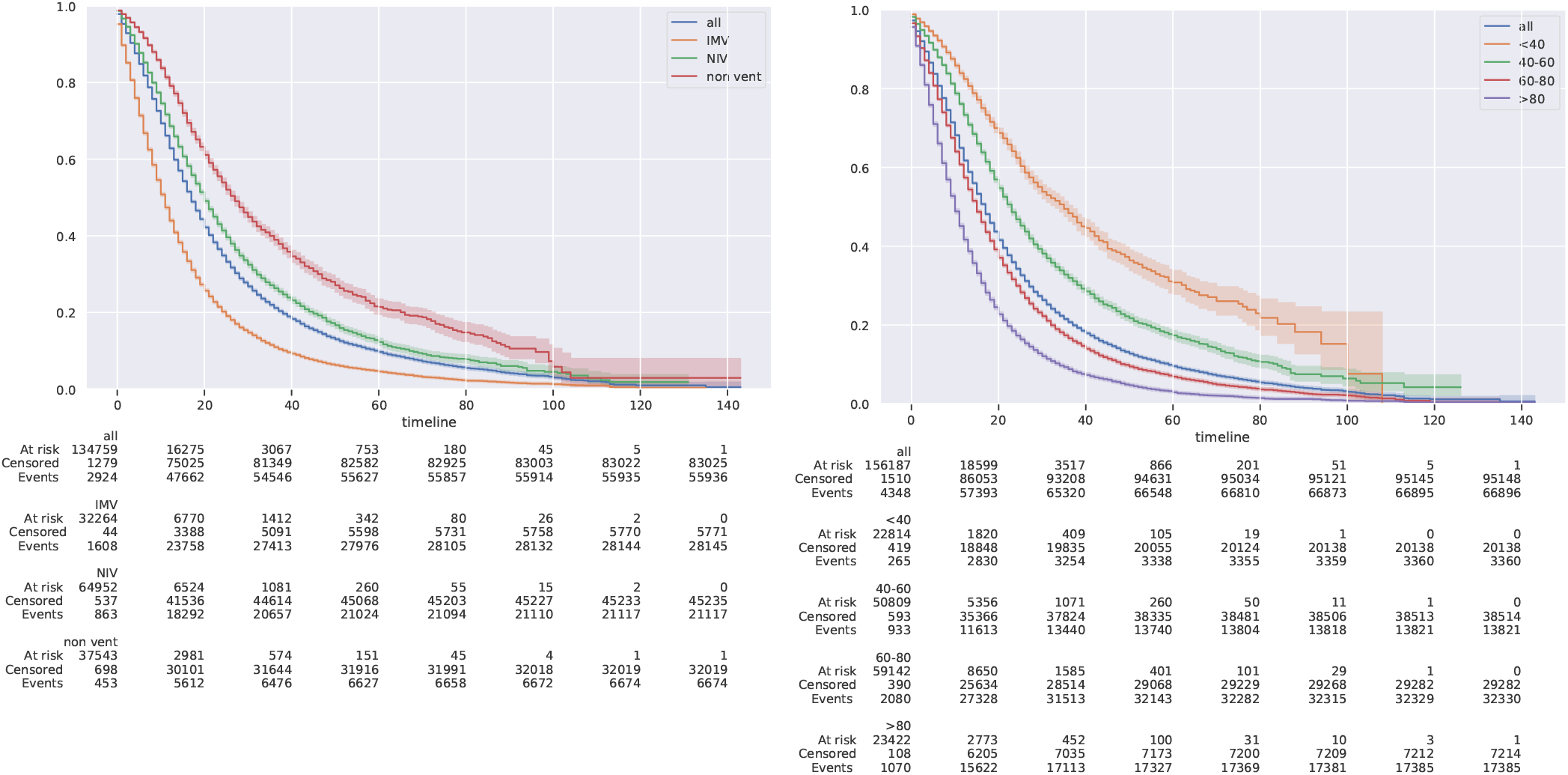
Kaplan-Meier survival curves by ventilation outcomes (n=138,962) (a) and age range (n=162,045) (b). IMV=Invasive Mechanical Ventilation, NIV=Non Invasive Ventilation.

### 3.3. Risk factors for mortality by subgroups

Hospitalized COVID-19 patients data was disaggregated according to ventilation requirement and age range, whose hazard ratios are shown in Figures 7 and 8, respectively (all data referring to such figures are presented in supplementary Tables S5-S11). In this case, a multiple Cox regression model was fitted for each subgroup, following the same variable selection strategy described before. Therefore, a different set of variables showed significant in each subgroup.

**Figure 7:**
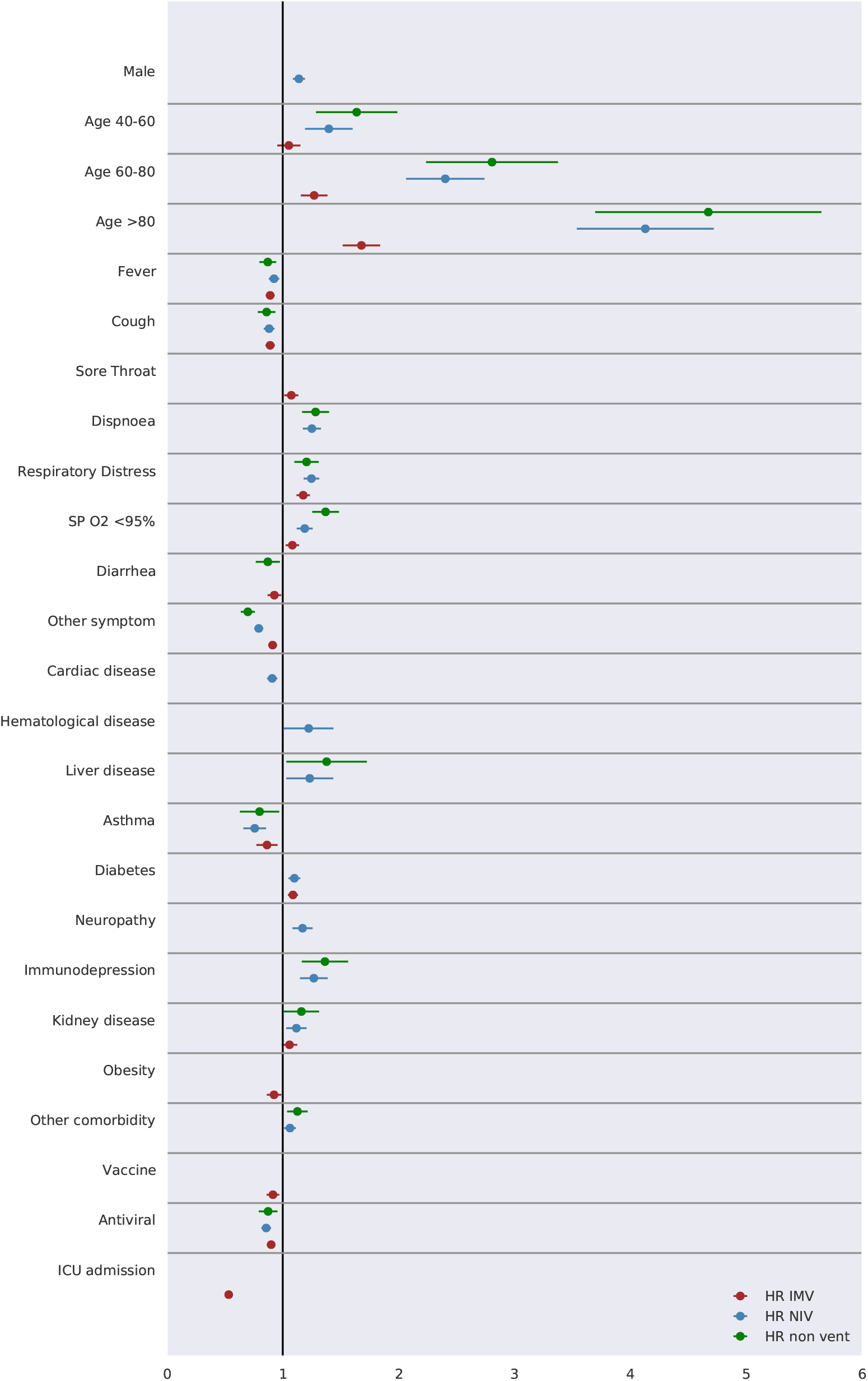
Mortality prognosis according to patient ventilation requirement in a multiple Cox regression model (95% CI) (see Tables S5-S7).

**Figure 8:**
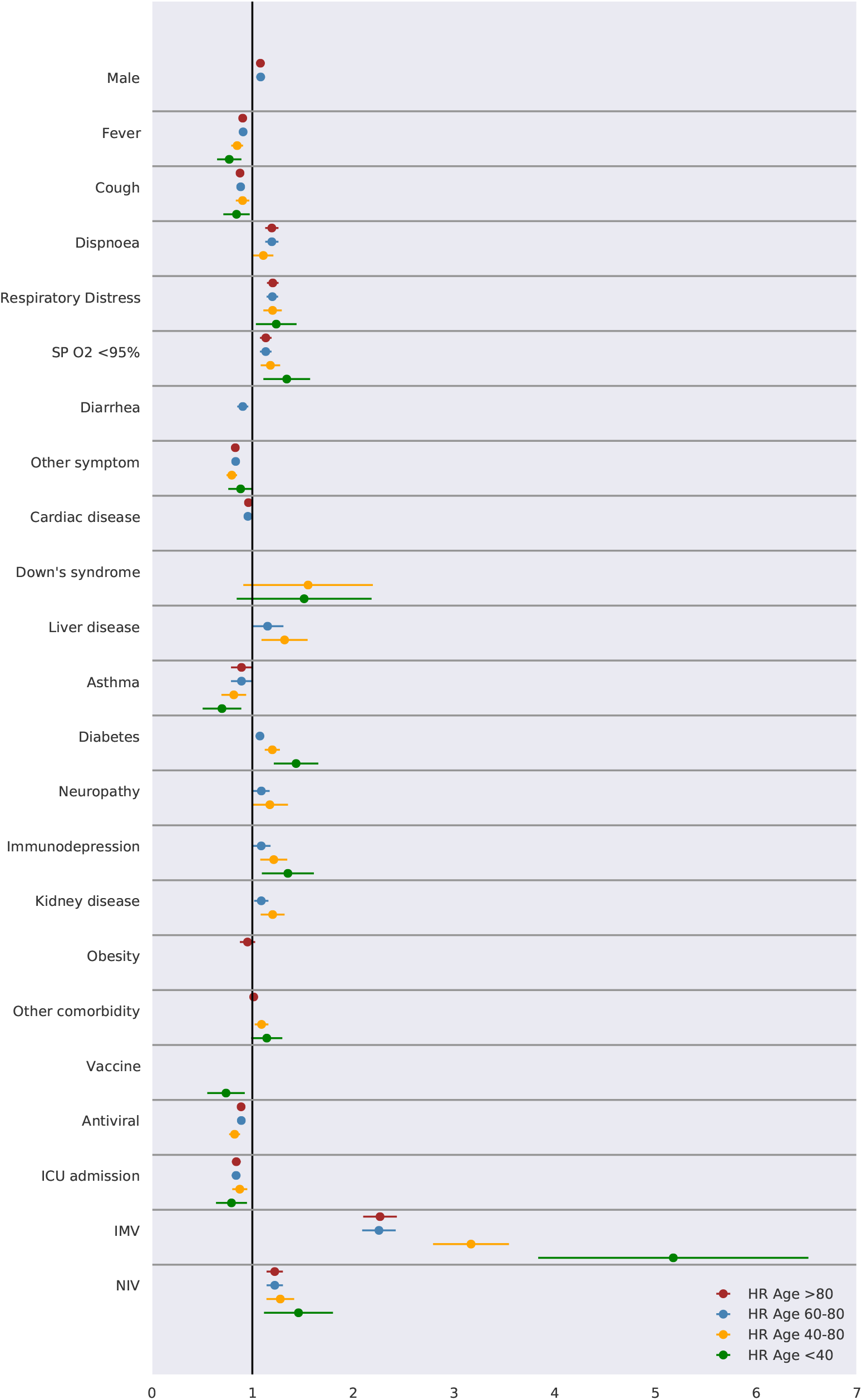
Mortality prognosis according to patient age range in a multiple Cox regression model (95% CI) (see Tables S8-S11).

According to Figure 7, older age is a risk factor independent of ventilation requirement or not. Clinical symptoms related to respiratory function were the most critical risk factors for NIV and non-ventilation patients. In addition, liver disease, immunosuppression and kidney disease were significantly associated with increased risk of mortality among NIV and non-ventilation patients. Diabetes presents a slight increase in the risk of mortality for patients with ventilation requirement (IMV and NIV). Influenza vaccine showed a small reduction in the risk of mortality only for patients that required IMV. On the other hand, the data suggests a small reduction in the risk of mortality for patients treated with Influenza antiviral regardless of the requirement for invasive ventilation.

The analysis of risk factors for different age subgroups unveiled interesting results (Figure 8). Respiratory distress and low oxygen saturation were a risk factor for all ages and dyspnoea for patients over 40 years old. Ventilation requirement (IMV or NIV) are mortality risk factors for all ages, but it should be noted that IMV is a relevant risk factor especially for patients *<*40 years.

Importantly, diabetes (HR=1.43; 95% CI, 1.23-1.67) and immunodepression (HR=1.35; 95% CI, 1.12-1.63) are also mortality risk factors especially for younger patients (*<*40 years). The disaggregation by age range showed an interesting result for the Influenza vaccine. There was a reduction in the mortality risk for vaccinated patients aged *<*40 years (HR=0.74; CI95%, 0.58-0.95). On the other hand, Influenza antiviral showed a reduced risk of mortality for patients ¿40 years.

In synthesis, hospitalized COVID-19 patients *<*40 years who are diabetic, immunosuppressed and requiring ventilatory support (IMV or NIV) are more vulnerable to the disease. On the other hand, older patients, male gender, presenting problems related to respiratory function and comorbidities such as liver disease, diabetes, neuropathy, immunodepression, kidney disease and requiring mechanical ventilation are at high mortality risk.

## 4. DISCUSSION

Using the major Brazilian database for COVID-19 cases registration, we built a comprehensive dataset containing demographic data, clinical characteristics and comorbidities of hospitalized patients in order to investigate the epidemiological and clinical profile of the study population and the main mortality risk factors. The analysis of mortality risk factors is relevant to define criteria for classifying the severity of the disease and to provide better care to those who may need ICU admission and ventilatory support.

The profile of Brazilian hospitalized patients who deceased is similar to hospitalized U.S. patients who died, whose characteristics were older age, male gender, immunosuppression, kidney disease, chronic lung disease, cardiovascular disease, neurologic disorders, and diabetes [18].

A meta-analysis including COVID-19 patients from different countries showed that age is a determining factor on mortality, especially above 60 years of age [19]. Although susceptibility to infection may be similar in different age ranges, studies suggest that susceptibility to symptomatic infection increases with age, possibly related to immunosenescence and unregulated immune response [20]. A recent study showed that although the fatality rate is higher in the elderly population, the lethality associated with hypertension and diabetes comorbidities is more pronounced in younger people [21]. Here, we also observed that younger brazilian patients with diabetes and immunosuppression comorbidities are also more vulnerable to COVID-19.

In addition to older age, we found that male gender is an independent risk factor for mortality in COVID-19 patients. Our data are in line with a large study that covered data from 23 European countries, which showed that the mortality rate of COVID-19 patients is significantly higher in male than in female, regardless of socioeconomic characteristics and health systems in these countries [22]. A study conducted with hospitalized COVID-19 patients using the same Brazilian database showed that the mortality rate in the North region is higher, and in Pardo and Black patients, at least partly due to non-ICU admission [23]. Our data demonstrated that the mortality rate is higher in the North/Northeast region and among brown color patients being in agreement with this study.

A study using the same database (SIVEP-gripe) covering the period from January 1st, 2020 to June 8th, 2020 found that COVID-19 patients who received a recent influenza vaccine (n=36,650) had 8% less chance to need intensive care, 18% less chance of requiring invasive ventilation and 17% less chance of death [24]. However, in our multiple Cox regression analysis (n=62,695 with vaccine status), we found no significant association between influenza vaccination and reduced risk of death, except for specific subgroups: patients under 40 years old and patients with IMV requirement. These differences may be due to our data extension (February 26 to August 10, 2020), a period when the majority of the population had been vaccinated in the 2020 campaign.

The rate of hospitalized COVID-19 patients with obesity was 11.76%, of whom 43.40% died (Table 2). The multiple regression analysis for obesity showed no significant association with mortality risk. Some studies have analyzed the association of obesity with poor prognosis for COVID-19 patients. Obese patients are at increased risk of developing severe illness [14]. The study presented by [25] showed a significant hazard ratio only for BMI ≥ 40Kg/m^2^ and, in another study, it was shown that BMI ≥ 35 kg/m^2^, in addition to increasing age and male gender, were independently associated with higher in-hospital mortality [26]. On the other hand, BMI *>*40 kg/m^2^, male gender and *<*60 years old were associated with higher mortality risk [27]. In addition to being a risk factor for severe COVID-19, high BMI was significantly associated with IMV requirement [28]. However, the Brazilian database does not discriminate overweight categories and BMI data is present for a few patients, so we were unable to perform categorized analysis.

The characterization of the profile of Brazilian patients admitted to ICU and requiring ventilation is relevant, as it can contribute to better care and reduced mortality. A recent review and meta-analysis involving COVID-19 patient data from several countries showed an ICU admission rate of 32%. The prevalence of mortality in the ICU was 39%, whereas in China this rate was higher, 42% [29]. Our analysis indicated that the events are more critical in Brazil, with 39.37% ICU admission and 62.40% mortality rate (Table 3), which may be related to infrastructure and availability of health professionals in public and private hospitals.

In accordance with some studies [25], [30], our data show that the invasive mechanical ventilation requirement among hospitalized COVID-19 patients is significant (24.41%), and with a extremely high mortality rate (82.98%) (Table 3). These data may be due to the high percentage of severely ill-hospitalized COVID-19 patients.

A meta-analysis study compiling data from several countries showed broad variability in ICU admission, IMV requirement and IMV mortality rates [31]. Brazilian COVID-19 patients ICU admission rate (39.37%) was similar a USA patients (35%). However, ICU mortality rate was extremely high (62.40%) compared to UK (33%), USA (29%), Italy (26%), China (24%), Spain (23%), France (15%), and Mexico (2%). Likewise, IMV patients mortality rate (82.98%) was higher than China (59%), UK (53%), USA (24%) and Mexico (4%). These high mortality rates may be due to late admission to the ICU and delay in the mechanical ventilation introduction, or even the need to change the care protocol.

A recent study showed that asthmatic patients are not at risk and do not develop severe SARS-COV-2 pneumonia compared to non-asthmatic patients [32]. In fact, we observed a low prevalence of asthmatic individuals (7.15%) among hospitalized COVID-19 patients, in agreement with the existing reports. Of note, the rate of cured asthmatics was twice that of those who died (Table 2) and asthma did not appear as a mortality risk factor (HR=0.82; 95% CI, 0.76-0.88, p*<*0.001). It is speculated that type 2 immune response and therapeutic drugs used for asthma may have a potential protective effect [33].

Highly prevalent comorbidities among hospitalized COVID-19 patients were cardiac disease and diabetes, which is in line with observations made in other populations [8], [12]. In our previous cohort study on outpatients from a Brazilian state (ES) [34], we found a prevalence of 18.55% for cardiac disease and 7.89% for diabetes. Mortality rates in this cohort were 50.05% and 31.82% for cardiac and diabetic patients, respectively. Here, the in-patient study showed a prevalence of 65.7% and 53.55% for cardiac disease and diabetes comorbidities, respectively (Table 2). Mortality rates for patients with such comorbidities are extremely high, around 50%. Therefore, these observations indicate that patients with cardiac disease and diabetes are subgroups that deserve special attention.

The main strength of this study is its scope, as it involved data from more than 162,000 hospitalized COVID-19 patients obtained from the major Brazilian Database. Thus, it was possible to outline the demographic and clinical profile of hospitalized COVID-19 patients who were cured and those who deceased. Moreover, risk factors for mortality by different subgroups were analysed.

Among the limitations of the study, we highlight the absence of data from biochemical tests of patients in SIVEP-GRIPE Database, which are relevant in prognostic studies. Another aspect is the notification data of only hospitalized COVID-19 patients, so that we were unable to trace the profile of outpatients. However, our previous study using a dataset from Espírito Santo state [34] can be used for comparative purposes, as it involved COVID-19 outpatients.

Taken together, our study provides a comprehensive overview of the epidemiological and clinical profile of Brazilian hospitalized COVID-19 patients and the analysis of the risk factors for mortality. We verified that older age and IMV requirement were the most significant risk factors for mortality, besides male gender and the presence of comorbidities. In the absence of vaccines or specific therapeutic drugs, prevention is the best strategy to protect people, specially this most vulnerable segment of the population.

The identification of groups at risk for severe COVID-19 is also important to establish priority groups for vaccination as soon as the vaccine is available, since the initial supply should be restricted. We believe this is the first comprehensive study that profiles COVID-19 patients in Brazil and highlights mortality risk factors during hospitalization.

## Data Availability

The data used in this work is publicly available.

## Acknowledgement

We would like to thank the Brazilian Ministry of Health for placing the COVID-19 database open access.

## Author Contributions

FSHS, NSHS and DLG performed the study design. FSHS, CMS and DLG extracted the data, built the dataset and analyzed all the data. FSHS and BDOB performed the statistical analysis. NSHS supported the literature review on COVID-19 and performed data comparisons. FSHS supervised the study. All authors contributed to the study, reviewed this article and approved the submitted version.

## Availability of data and material

The data used in this work is publicly available in https://opendatasus.saude.gov.br/dataset/bd-srag-2020

## Supplementary Material

**TABLE S1:** Demographic data of the study population (n=44,128)

|  | all n(%) | cure n(%) | death n(%) | lr<sup>a</sup>(%) |
| --- | --- | --- | --- | --- |
| Region | 44128 (100.00) | 24034 (54.46) | 20094 (45.54) |  |
| South | 6147 (13.93) | 3740 (15.56) | 2407 (11.98) | 39.16 |
| Southeast | 24321 (55.11) | 13842 (57.59) | 10479 (52.15) | 43.09 |
| Midwest | 3676 (8.33) | 2041 (8.49) | 1635 (8.14) | 44.48 |
| Northeast | 7571 (17.16) | 3410 (14.19) | 4161 (20.71) | 54.96 |
| North | 2413 (5.47) | 1001 (4.16) | 1412 (7.03) | 58.52 |
| Age Range<sup>b</sup> |  |  |  |  |
| 0-4 | 233 (0.53) | 188 (0.78) | 45 (0.22) | 19.31 |
| 5-9 | 91 (0.21) | 82 (0.34) | 9 (0.04) | 9.89 |
| 10-19 | 251 (0.57) | 203 (0.84) | 48 (0.24) | 19.12 |
| 20-29 | 942 (2.13) | 749 (3.12) | 193 (0.96) | 20.49 |
| 30-39 | 2661 (6.03) | 2098 (8.73) | 563 (2.80) | 21.16 |
| 40-49 | 4854 (11.00) | 3619 (15.06) | 1235 (6.15) | 25.44 |
| 50-59 | 7881 (17.86) | 5264 (21.90) | 2617 (13.02) | 33.21 |
| 60-69 | 10193 (23.10) | 5565 (23.15) | 4628 (23.03) | 45.40 |
| 70-79 | 9210 (20.87) | 3851 (16.02) | 5359 (26.67) | 58.19 |
| 80-89 | 6139 (13.91) | 1984 (8.25) | 4155 (20.68) | 67.68 |
| 90+ | 1673 (3.79) | 431 (1.79) | 1242 (6.18) | 74.24 |
| Gender |  |  |  |  |
| male | 23895 (54.15) | 12558 (52.25) | 11337 (56.42) | 47.45 |
| female | 20233 (45.85) | 11476 (47.75) | 8757 (43.58) | 43.28 |
| Race | 34505 (100.00) | 18307 (53.06) | 16198 (46.94) |  |
| white | 18329 (53.12) | 10304 (56.28) | 8025 (49.54) | 43.78 |
| black | 2337 (6.77) | 1213 (6.63) | 1124 (6.94) | 48.10 |
| asian | 467 (1.35) | 235 (1.28) | 232 (1.43) | 49.68 |
| brown | 13315 (38.59) | 6529 (35.66) | 6786 (41.89) | 50.97 |
| indigenous | 57 (0.17) | 26 (0.14) | 31 (0.19) | 54.39 |
| Education<sup>c</sup> | 19376 (100.00) | 10750 (55.48) | 8626 (44.52) |  |
| illiterate | 1386 (7.15) | 510 (4.74) | 876 (10.16) | 63.20 |
| ES-1 | 5793 (29.90) | 2675 (24.88) | 3118 (36.15) | 53.82 |
| ES-2 | 3870 (19.97) | 2096 (19.50) | 1774 (20.57) | 45.84 |
| HS | 5605 (28.93) | 3509 (32.64) | 2096 (24.30) | 37.40 |
| HE | 2549 (13.16) | 1822 (16.95) | 727 (8.43) | 28.52 |
| NA | 173 (0.89) | 138 (1.28) | 35 (0.41) | 20.23 |

a. lethality rate
b. in years
c. ES-1= Elementary School 1; ES-2= Elementary School 2; HS= High School; HE= Higher Education, NA= Not Applicable (age<7)

**TABLE S2:** Clinical data of the study population (n=44,128)

|  | all n(%) | cure n(%) | death n(%) |
| --- | --- | --- | --- |
| Symptom |  |  |  |
| Fever (n=44128) | 29824 (67.59) | 16800 (56.33) | 13024 (43.67) |
| Cough (n=44128) | 33008 (74.80) | 18762 (56.84) | 14246 (43.16) |
| Sore Throat (n=44128) | 7601 (17.22) | 4522 (59.49) | 3079 (40.51) |
| Dispnoea (n=44128) | 33053 (74.90) | 16675 (50.45) | 16378 (49.55) |
| Respiratory Distress (n=44128) | 27863 (63.14) | 13518 (48.52) | 14345 (51.48) |
| SP O2 < 95%<sup>a</sup> (n=44128) | 29358 (66.53) | 13882 (47.29) | 15476 (52.71) |
| Diarrhea (n=44128) | 6338 (14.36) | 3940 (62.16) | 2398 (37.84) |
| Vomit (n=44128) | 3766 (8.53) | 2273 (60.36) | 1493 (39.64) |
| Other (n=44128) | 17488 (39.63) | 10889 (62.27) | 6599 (37.73) |
| Comorbidity |  |  |  |
| Cardiac disease (n=44128) | 22957 (52.02) | 11827 (51.52) | 11130 (48.48) |
| Hematological disease (n=44128) | 650 (1.47) | 305 (46.92) | 345 (53.08) |
| Down's syndrome (n=44128) | 203 (0.46) | 108 (53.20) | 95 (46.80) |
| Liver disease (n=44128) | 733 (1.66) | 298 (40.65) | 435 (59.35) |
| Asthma (n=44128) | 2118 (4.80) | 1425 (67.28) | 693 (32.72) |
| Diabetes (n=44128) | 17573 (39.82) | 8825 (50.22) | 8748 (49.78) |
| Neuropathy (n=44128) | 2866 (6.49) | 1051 (36.67) | 1815 (63.33) |
| Pneumopathy (n=44128) | 2788 (6.32) | 1101 (39.49) | 1687 (60.51) |
| Immunodepression (n=44128) | 2343 (5.31) | 1046 (44.64) | 1297 (55.36) |
| Kidney disease (n=44128) | 3227 (7.31) | 1176 (36.44) | 2051 (63.56) |
| Obesity (n=44128) | 3633 (8.23) | 2119 (58.33) | 1514 (41.67) |
| Other (n=44128) | 20081 (45.51) | 10548 (52.53) | 9533 (47.47) |

a. oxygen saturation

**TABLE S3:** Additional information of the study population (n=44,128)

|  | all n(%) | cure n(%) | death n(%) |
| --- | --- | --- | --- |
| Vaccine | 8234 (18.66) | 4729 (57.43) | 3505 (42.57) |
| Antiviral | 14732 (33.38) | 8161 (55.40) | 6571 (44.60) |
| ICU admission | 18590 (42.13) | 6222 (33.47) | 12368 (66.53) |
| Ventilation | 32940 (74.65) | 15395 (46.74) | 17545 (53.26) |
| IMV<sup>a</sup> | 11747 (26.62) | 1798 (15.31) | 9949 (84.69) |
| NIV<sup>b</sup> | 21193 (48.03) | 13597 (64.16) | 7596 (35.84) |

a. Invasive Mechanical Ventilation
b. Non Invasive Ventilation

**TABLE S4:** Risk factors in fatal outcome using a simple Cox regression model (95% CI)

| Variable<sup>a</sup> | HR | CI 95% | p-value |
| --- | --- | --- | --- |
| Male | 1.03 | (1.00-1.06) | 0.044 |
| Age 40-60 | 0.62 | (0.60-0.64) | <0.001 |
| Age 60-80 | 1.10 | (1.07-1.13) | <0.001 |
| Age >80 | 1.80 | (1.74-1.85) | <0.001 |
| Fever | 0.83 | (0.81-0.85) | <0.001 |
| Cough | 0.83 | (0.80-0.85) | <0.001 |
| Sore Throat | 0.92 | (0.89-0.96) | <0.001 |
| Dispnoea | 1.44 | (1.39-1.50) | <0.001 |
| Respiratory Distress | 1.46 | (1.41-1.50) | <0.001 |
| SP O2 <95% | 1.53 | (1.48-1.58) | <0.001 |
| Diarrhea | 0.84 | (0.81-0.88) | <0.001 |
| Vomit | 0.87 | (0.82-0.91) | <0.001 |
| Other symptom | 0.76 | (0.74-0.79) | <0.001 |
| Cardiac disease | 1.10 | (1.07-1.13) | <0.001 |
| Hematological disease | 1.05 | (0.94-1.17) | 0.389 |
| Down's syndrome | 1.04 | (0.85-1.27) | 0.693 |
| Liver disease | 1.23 | (1.12-1.35) | <0.001 |
| Asthma | 0.74 | (0.68-0.79) | <0.001 |
| Diabetes | 1.14 | (1.11-1.18) | <0.001 |
| Neuropathy | 1.29 | (1.23-1.35) | <0.001 |
| Pneumopathy | 1.20 | (1.14-1.26) | <0.001 |
| Immunodepression | 1.07 | (1.01-1.13) | 0.018 |
| Kidney disease | 1.21 | (1.16-1.27) | <0.001 |
| Obesity | 0.88 | (0.83-0.93) | <0.001 |
| Other comorbidity | 1.03 | (1.00-1.06) | 0.055 |
| Vaccine | 0.96 | (0.92-0.99) | 0.015 |
| Antiviral | 0.89 | (0.87-0.92) | <0.001 |
| ICU admission | 1.37 | (1.33-1.41) | <0.001 |
| IMV | 2.01 | (1.95-2.06) | <0.001 |
| NIV | 0.74 | (0.72-0.76) | <0.001 |

a. see Table S2; n=44,128 patients with full symptom/comorbidity information

**TABLE S5:** Risk factors in fatal outcome using a multiple Cox regression model (95% CI) for the IMV subgroup

| Variable | HR | CI 95% | p value |
| --- | --- | --- | --- |
| Age 40-60 | 1.05 | (0.96-1.16) | 0.289 |
| Age 60-80 | 1.27 | (1.16-1.39) | <0.001 |
| Age >80 | 1.68 | (1.53-1.85) | <0.001 |
| Fever | 0.89 | (0.85-0.93) | <0.001 |
| Cough | 0.89 | (0.85-0.93) | <0.001 |
| Sore Throat | 1.07 | (1.01-1.14) | 0.014 |
| Respiratory Distress | 1.18 | (1.12-1.24) | <0.001 |
| SP O2 <95% | 1.08 | (1.03-1.14) | <0.005 |
| Diarrhea | 0.93 | (0.87-0.99) | 0.020 |
| Other symptom | 0.91 | (0.88-0.95) | <0.001 |
| Asthma | 0.86 | (0.78-0.96) | 0.007 |
| Diabetes | 1.09 | (1.05-1.13) | <0.001 |
| Kidney disease | 1.06 | (0.99-1.13) | 0.077 |
| Obesity | 0.93 | (0.86-0.99) | 0.024 |
| Vaccine | 0.92 | (0.86-0.97) | <0.005 |
| Antiviral | 0.90 | (0.86-0.94) | <0.001 |
| ICU admission | 0.53 | (0.50-0.57) | <0.001 |

**TABLE S6:** Risk factors in fatal outcome using a multiple Cox regression model (95% CI) for the NIV subgroup

| Variable | HR | CI 95% | p value |
| --- | --- | --- | --- |
| Male | 1.14 | (1.09-1.19) | <0.001 |
| Age 40-60 | 1.40 | (1.21-1.62) | <0.001 |
| Age 60-80 | 2.40 | (2.09-2.76) | <0.001 |
| Age >80 | 4.13 | (3.58-4.76) | <0.001 |
| Fever | 0.92 | (0.88-0.97) | <0.005 |
| Cough | 0.88 | (0.84-0.93) | <0.001 |
| Dispnoea | 1.25 | (1.18-1.33) | <0.001 |
| Respiratory Distress | 1.25 | (1.18-1.32) | <0.001 |
| SP O2 <95% | 1.19 | (1.12-1.26) | <0.001 |
| Other symptom | 0.79 | (0.76-0.83) | <0.001 |
| Cardiac disease | 0.91 | (0.87-0.95) | <0.001 |
| Hematological disease | 1.22 | (1.03-1.46) | 0.023 |
| Liver disease | 1.23 | (1.05-1.45) | 0.012 |
| Asthma | 0.76 | (0.67-0.86) | <0.001 |
| Diabetes | 1.10 | (1.05-1.15) | <0.001 |
| Neuropathy | 1.17 | (1.09-1.26) | <0.001 |
| Immunodepression | 1.27 | (1.15-1.39) | <0.001 |
| Kidney disease | 1.12 | (1.03-1.21) | 0.006 |
| Other comorbidity | 1.06 | (1.01-1.12) | 0.013 |
| Antiviral | 0.86 | (0.82-0.90) | <0.001 |

**TABLE S7:** Risk factors in fatal outcome using a multiple Cox regression model (95% CI) for the non ventilation subgroup

| Variable | HR | CI 95% | p value |
| --- | --- | --- | --- |
| Age 40-60 | 1.64 | (1.32-2.03) | <0.001 |
| Age 60-80 | 2.81 | (2.29-3.43) | <0.001 |
| Age >80 | 4.67 | (3.80-5.75) | <0.001 |
| Fever | 0.87 | (0.80-0.95) | <0.005 |
| Cough | 0.86 | (0.79-0.94) | <0.001 |
| Dispnoea | 1.28 | (1.17-1.41) | <0.001 |
| Respiratory Distress | 1.21 | (1.10-1.32) | <0.001 |
| SP O2 <95% | 1.37 | (1.26-1.49) | <0.001 |
| Diarrhea | 0.87 | (0.77-0.98) | 0.023 |
| Other symptom | 0.70 | (0.64-0.76) | <0.001 |
| Liver disease | 1.38 | (1.07-1.77) | 0.012 |
| Asthma | 0.80 | (0.65-0.99) | 0.038 |
| Immunodepression | 1.36 | (1.18-1.58) | <0.001 |
| Kidney disease | 1.16 | (1.02-1.32) | 0.026 |
| Other comorbidity | 1.13 | (1.04-1.22) | <0.005 |
| Antiviral | 0.87 | (0.80-0.96) | <0.005 |

**TABLE S8:** Risk factors in fatal outcome using a multiple Cox regression model (95% CI) for the Age *<*40 subgroup

| Variable | HR | CI 95% | p value |
| --- | --- | --- | --- |
| Fever | 0.77 | (0.66-0.90) | <0.005 |
| Cough | 0.84 | (0.72-0.98) | 0.031 |
| Respiratory Distress | 1.24 | (1.05-1.46) | 0.010 |
| SP O2 <95% | 1.34 | (1.13-1.59) | <0.001 |
| Other symptom | 0.88 | (0.77-1.01) | 0.079 |
| Down's syndrome | 1.51 | (0.99-2.32) | 0.058 |
| Asthma | 0.70 | (0.53-0.92) | 0.010 |
| Diabetes | 1.43 | (1.23-1.67) | <0.001 |
| Immunodepression | 1.35 | (1.12-1.63) | <0.005 |
| Other comorbidity | 1.14 | (1.00-1.31) | 0.051 |
| Vaccine | 0.74 | (0.58-0.95) | 0.017 |
| ICU admission | 0.79 | (0.65-0.96) | 0.018 |
| IMV | 5.18 | (4.01-6.69) | <0.001 |
| NIV | 1.46 | (1.16-1.84) | <0.005 |

**TABLE S9:**
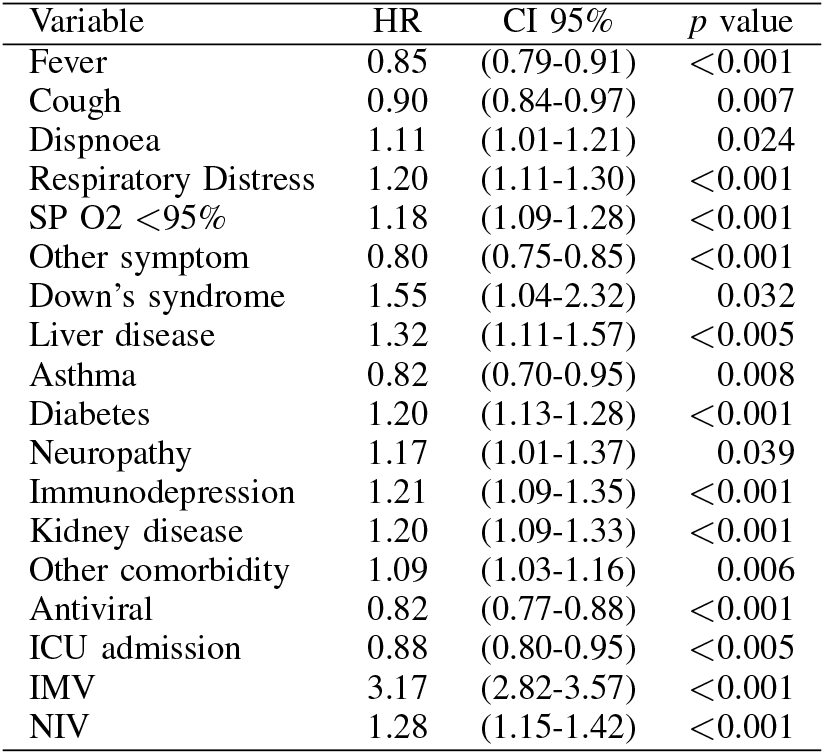
Risk factors in fatal outcome using a multiple Cox regression model (95% CI) for the Age 40-60 subgroup

| Variable | HR | CI 95% | *p* value |
| --- | --- | --- | --- |
| Fever | 0.85 | (0.79-0.91) | <0.001 |
| Cough | 0.90 | (0.84-0.97) | 0.007 |
| Dispnoea | 1.11 | (1.01-1.21) | 0.024 |
| Respiratory Distress | 1.20 | (1.11-1.30) | <0.001 |
| SP O2 <95% | 1.18 | (1.09-1.28) | <0.001 |
| Other symptom | 0.80 | (0.75-0.85) | <0.001 |
| Down's syndrome | 1.55 | (1.04-2.32) | 0.032 |
| Liver disease | 1.32 | (1.11-1.57) | <0.005 |
| Asthma | 0.82 | (0.70-0.95) | 0.008 |
| Diabetes | 1.20 | (1.13-1.28) | <0.001 |
| Neuropathy | 1.17 | (1.01-1.37) | 0.039 |
| Immunodepression | 1.21 | (1.09-1.35) | <0.001 |
| Kidney disease | 1.20 | (1.09-1.33) | <0.001 |
| Other comorbidity | 1.09 | (1.03-1.16) | 0.006 |
| Antiviral | 0.82 | (0.77-0.88) | <0.001 |
| ICU admission | 0.88 | (0.80-0.95) | <0.005 |
| IMV | 3.17 | (2.82-3.57) | <0.001 |
| NIV | 1.28 | (1.15-1.42) | <0.001 |

**TABLE S10:** Risk factors in fatal outcome using a multiple Cox regression model (95% CI) for the Age 60-80 subgroup

| Variable | HR | CI 95% | *p* value |
| --- | --- | --- | --- |
| Male | 1.08 | (1.04-1.13) | <0.001 |
| Fever | 0.91 | (0.87-0.95) | <0.001 |
| Cough | 0.88 | (0.85-0.92) | <0.001 |
| Dispnoea | 1.19 | (1.13-1.26) | <0.001 |
| Respiratory Distress | 1.20 | (1.14-1.26) | <0.001 |
| SP O2 <95% | 1.13 | (1.08-1.19) | <0.001 |
| Diarrhea | 0.90 | (0.85-0.96) | <0.005 |
| Other symptom | 0.83 | (0.80-0.87) | <0.001 |
| Cardiac disease | 0.96 | (0.92-0.99) | 0.027 |
| Liver disease | 1.15 | (1.01-1.32) | 0.042 |
| Asthma | 0.89 | (0.79-1.00) | 0.055 |
| Diabetes | 1.08 | (1.03-1.12) | <0.001 |
| Neuropathy | 1.09 | (1.01-1.17) | 0.026 |
| Immunodepression | 1.09 | (1.00-1.18) | 0.050 |
| Kidney disease | 1.09 | (1.02-1.16) | 0.010 |
| Antiviral | 0.89 | (0.85-0.93) | <0.001 |
| ICU admission | 0.84 | (0.80-0.88) | <0.001 |
| IMV | 2.26 | (2.10-2.43) | <0.001 |
| NIV | 1.22 | (1.14-1.31) | <0.001 |

**TABLE S11:** Risk factors in fatal outcome using a multiple Cox regression model (95% CI) for the Age *>*80 subgroup

| Variable | HR | CI 95% | *p* value |
| --- | --- | --- | --- |
| Male | 1.08 | (1.04-1.12) | <0.001 |
| Fever | 0.90 | (0.87-0.94) | <0.001 |
| Cough | 0.88 | (0.84-0.92) | <0.001 |
| Dispnoea | 1.19 | (1.13-1.26) | <0.001 |
| Respiratory Distress | 1.20 | (1.15-1.26) | <0.001 |
| SP O2 <95% | 1.13 | (1.08-1.19) | <0.001 |
| Other symptom | 0.83 | (0.80-0.87) | <0.001 |
| Cardiac disease | 0.96 | (0.92-1.00) | 0.063 |
| Asthma | 0.89 | (0.79-1.00) | 0.056 |
| Obesity | 0.95 | (0.88-1.03) | 0.227 |
| Other comorbidity | 1.01 | (0.97-1.06) | 0.523 |
| Antiviral | 0.89 | (0.85-0.93) | <0.001 |
| ICU admission | 0.84 | (0.80-0.89) | <0.001 |
| IMV | 2.27 | (2.11-2.44) | <0.001 |
| NIV | 1.22 | (1.14-1.31) | <0.001 |

## Footnotes

1 https://opendatasus.saude.gov.br/dataset/bd-srag-2020

2 The algorithm minimizes − 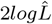, where 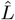 stands for the maximum likelihood of a variable.

3 Age categories count as one variable.

